# Definition of a new blood cell count (BCT) score for early survival prediction for non-small cell lung cancer patients treated with atezolizumab: Integrated analysis of 4 multicenter clinical trials

**DOI:** 10.1101/2021.08.28.21262770

**Authors:** Jian-Guo Zhou, Ada Hang-Heng Wong, Haitao Wang, Su-Han Jin, Fangya Tan, Yu-Zhong Chen, Si-Si He, Gang Shen, Benjamin Frey, Rainer Fietkau, Markus Hecht, Shamus R. Carr, Ruihong Wang, Bo Shen, David S Schrump, Hu Ma, Udo S. Gaipl

## Abstract

**Introduction:** Immune checkpoint inhibitor (ICI) therapy is a major breakthrough in non-small cell lung cancer (NSCLC) treatment. However, valid predictive biomarkers are lacking. Blood cell count test (BCT) provides a direct quantification of various types of immune cells (ICs) to reveal the immune landscape to predict ICI treatment.

**Methods:** This study analyzed four international, multi-center clinical trials (OAK, BIRCH, POPLAR and FIR trials) to conduct post-hoc analyses of NSCLC patients undergoing atezolizumab (anti-PD-L1) single-agent treatment (n = 1,479) or docetaxel single-agent treatment (n = 707). BCT was conducted at three timepoints: pre-treatment (T1), the first day of treatment cycle 3 (T2), and first day of treatment cycle 5 (T3). Univariate and multivariate Cox regression analyses were conducted to identify early BCT biomarkers to predict atezolizumab treatment outcomes in NSCLC patients.

**Results:** The BCT biomarkers of neutrophil-to-lymphocyte ratio (NLR) and platelet-to-lymphocyte ratio (PLR) at timepoint T3 and neutrophil-to-monocyte ratio (NMR) at timepoint T2 were identified as strong predictive biomarkers for atezolizumab (Ate)-treated NSCLC patients in comparison to docetaxel (Dtx)-treated patients regarding overall survival (OS) (BCTscore low-risk: HR _Ate vs Dtx_ = 1.54 (95% CI: 1.04-2.27), *P* = 0.036; high-risk: HR _Ate vs Dtx_ = 0.84 (95% CI: 0.62-1.12), *P* = 0.236). This identified BCTscore model showed better OS AUC in the OAK (AUC_12month_=0.696), BIRCH (AUC_12month_=0.672) and POPLAR+FIR studies (AUC_12month_=0.727) than that of each of the three single BCT biomarkers.

**Conclusion:** The BCTscore model is a valid predictive and prognostic biomarker for atezolizumab-treated NSCLC patients.

## Background

Non-small cell lung cancer (NSCLC) accounts for 84% of all lung cancer incidence, roughly accounting for 235,170 new cases in the U.S. in 2021 ^1^. Therapy for advanced NSCLC can include chemotherapy, radiotherapy, or tyrosine kinase inhibitors (TKIs) as first-line therapy for patients carrying genetic mutations in the genes of *EGFR, ALK, ROS1* and *NTRK* ^2^. However, for patients without TKI-targeted mutations, safe and effective therapeutic options were limited. With the development of immune checkpoint inhibitors (ICIs) this has changed. ICIs were developed against programmed cell death ligand 1 (PD-L1), and the immune suppressive receptors programmed cell death 1 (PD-1) and cytotoxic T lymphocyte-associated antigen 4 (CTLA-4), being present on cytotoxic T cells ^2^.

These therapies have improved NSCLC treatment outcomes in patients with advanced disease ^3-5^. However, without implementation of patient selection by predictive or prognostic biomarkers, no significant PFS or OS improvement by ICI therapy, as compared to chemotherapy, are observed ^6^. PD-L1 expression and tumor mutational burden (TMB) are often recommended for patient selection before treatment, but contrasting results are seen in clinical trials involving atezolizumab ^3, 4, 7^ and nivolumab ^6^.

Recent studies propose that chromosome instability, tumor microsatellite instability, and T-cell surface markers such as PD-1^†^, CD38 and CD39^‡^, or CD8^8^ might serve as prognostic and predictive biomarkers for ICI therapy ^9, 10^. However, genetic biomarkers require tumor biopsy samples which is invasive and limits longitudinal analysis for continuous disease monitoring. Hence, liquid biopsy-based biomarkers are coming more and more in the focus ^11-15^.

Blood cell count test (BCT) is a routine, regularly performed blood test conducted before and during treatment. BCT provides a direct overview of the immune landscape based on the counts of various types of immune cells (ICs). For instance, high pre-treatment neutrophil-to-lymphocyte ratio (NLR) and platelet-to-lymphocyte ratio (PLR) correlated with poor survival outcomes in NSCLC patients treated with ICIs, regardless of TMB ^16-18^. However, a limitation of most of the published studies are either small cohorts or analysis of multiple ICI therapies in diverse clinical settings which may compromise the validity of findings. Consequently, this study focuses on survival data obtained from four international, multi-center clinical trials to conduct post-hoc analysis of NSCLC patients undergoing atezolizumab (Ate) single-agent treatment, while docetaxel (Dtx) single-agent treatment served as control. BCT was conducted at three timepoints: baseline (T1), 6 weeks on-treatment (T2), and 12 weeks on-treatment (T3). The overarching goal was to identify a BCTscore as a biomarker that may predict ICI efficacy in NSCLC.

## Methods

### Study cohort

Pseudonymized individual participant data from the single-arm phase II studies FIR (NCT01846416) ^19^ and BIRCH (NCT02031458) ^20^, and the two-arm randomized controlled trials (RCTs) POPLAR phase II study (NCT01903993) ^3^ and OAK phase III study (NCT02008227) ^4^ were provided by Genentech Inc. and accessed through the secure Vivli online platform. Raw data were extracted and compared with the available published data to ensure accuracy. Secondary analysis of the trial data was deemed to be of negligible risk and was approved by the Institutional Review Board of the Second Affiliated Hospital, Zunyi Medical University (No.YXLL(KY-R)-2021-010). Deidentified data were accessed according to Roche’s policy and process for Vivli. Data analyses were conducted from March 2, 2021, to June 30, 2021.

A total of 2,316 patients were included from the four clinical trials, and after exclusion of untreated patients and patients without pre-treatment BCT, 1,479 and 707 advanced NSCLC patients undergoing atezolizumab and docetaxel treatment, respectively, were included in this study (Figure 1). Of note, atezolizumab was administered either as first-line or second-line therapy after failure of prior chemotherapy in the four trials used in this study. Atezolizumab and docetaxel were both administered every 3 weeks in the two-arm RCTs POPLAR and OAK. BCT was obtained at three timepoints: pre-treatment baseline (T1), 6 weeks on-treatment (T2), and 12 weeks on-treatment (T3). Baseline was defined as within 28 days prior to the start of treatment. Timepoints T2 and T3 corresponded to the first day of treatment cycles 3 and 5, respectively.

**Figure 1.**
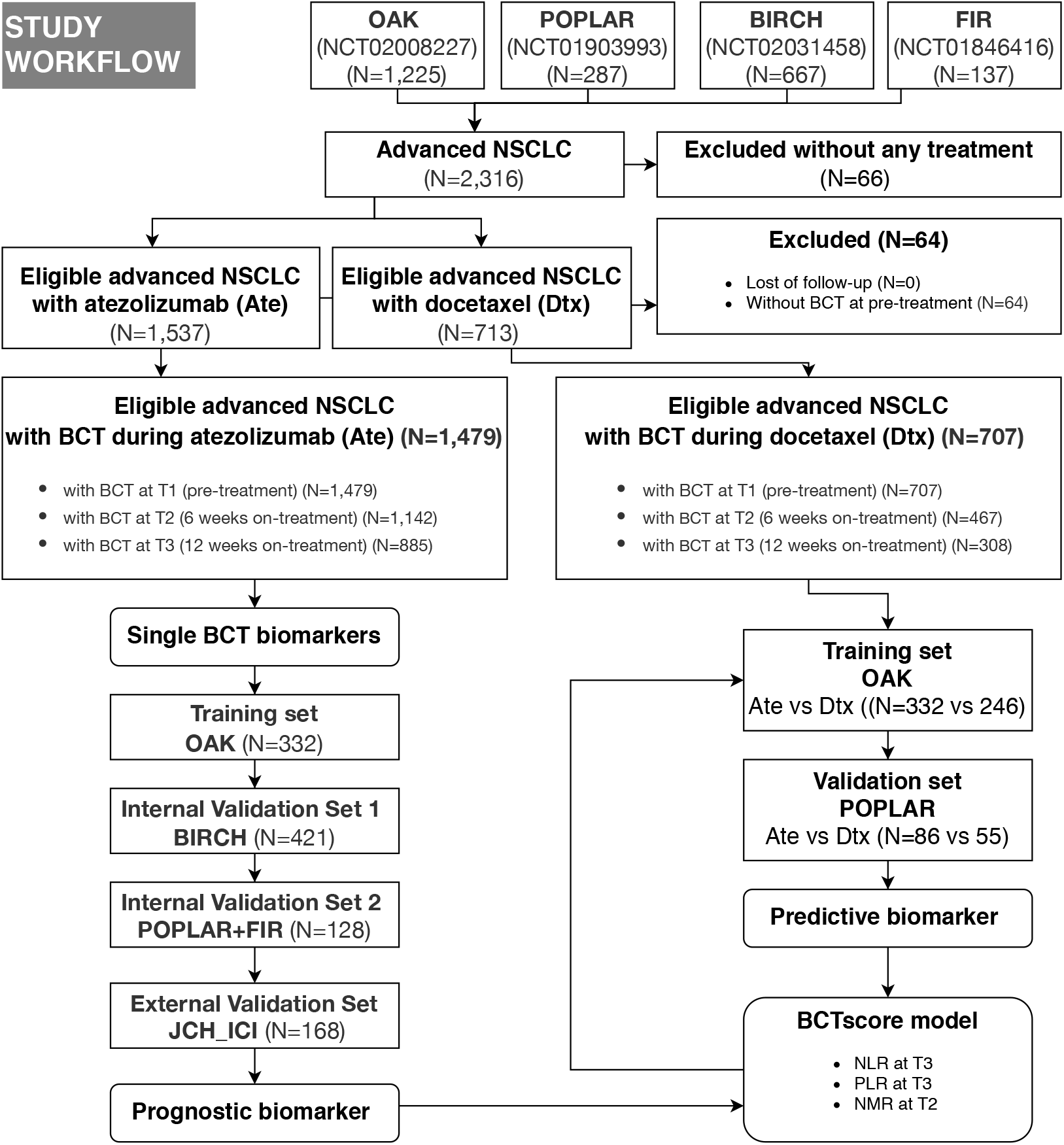
Flow chart demonstrating the patient cohorts of the indicated trials and the approach for the development of a blood cell count test (BCT)-based score (BCTscore). The internal cohorts are from four international, multicenter studies (OAK, POPLAR, BIRCH, and FIR), whereas the JCH_ICI cohort was used as an external validation cohort. Ate: atezelizumab; Dtx: docetaxel.

We then validated our results with an external cohort (JCH_ICI) containing 168 advanced or relapsed NSCLC patients who received anti-PD-1-based combination therapy and had an early BCT at the Affiliated Cancer Hospital of Nanjing Medical University, China, from August 2018 to December 2019 ^21^. In this cohort, patients received intravenous administration of combination therapy containing pembrolizumab, sintilimab, or toripalimab once every 3 weeks. Combination chemotherapy included platinum doublet chemotherapy in conjunction with pemetrexed, docetaxel, paclitaxel/nab-paclitaxel, gemcitabine, or bevacizumab.

### Predictor and treatment outcome definitions

The definitions of OS, PFS, clinical benefit (CB) and objective response rate (ORR) were detailed in each trial ^3, 4, 19, 20^. In this study, OS was used as the primary endpoint, whereas PFS according to Response Evaluation Criteria in Solid Tumors (RECIST version 1.1), CB, and ORR were used as secondary endpoints. The BCT biomarkers of NLR, PLR, NMR, and lymphocyte-to-monocyte ratio (LMR) were calculated by dividing absolute cell counts of corresponding ICs acquired from BCT at all timepoints, as described above.

### Statistical analysis

Associations between BCT biomarkers and OS or PFS were calculated by the Cox proportional hazards regression model and reported as the mean of hazard ratio (HR) with two-sided 95% confidence interval (CI) and *p*-value as calculated by the Wald test. The Kaplan-Meier method was used to estimate median OS and PFS between risk groups with a stratified log-rank test at the two-sided significance level. Survival analysis was performed by the *survival* (V.3.2-11) and *survminer* (V.0.4.9) packages. To analyze the degree of discrimination of biomarkers, we performed time-dependent receiver-operating characteristic (ROC) analysis and calculated the area under curve (AUC) for the indicated survival outcomes by the *timeROC* (V.0.3) and *pROC* (V.1.17.0.1) packages. Comparisons of CB, ORR, or clinical factors between the specified groups were calculated by the generalized linear model (GLM) to report relative risk (RR) with 95% CI, and *p*-value as calculated by the Pearson’s χ2-test or Fisher’s exact test.

Comparisons of BCT biomarkers between the treatment groups or different time points of the same treatment group were performed using Wilcoxon signed-rank test. All statistical analyses were carried out in R V.3.6.1 (R Foundation for Statistical Computing). *P* ≤ 0.050 was considered to be statistically significant. All analyses were univariate except for the multivariate Cox analyses. In multivariate analysis, the BCT biomarker(s) and the clinical factors of sex (male / female), age, race (white / Asian / other), Eastern Cooperative Oncology Group performance status (ECOG PS), metastasis, and pre-treatment PD-L1 (high ≥1% / low <1%; except for the BIRCH study, high ≥5% / low <5%) were included; however, the additional biomarkers of body mass index (BMI) and smoker (never / previous / current) were insignificant as assessed by univariate Cox analysis and were hence removed from the analyses.

## Results

### Identification of BCT biomarkers related to treatment outcomes of patients treated with atezolizumab but not of those treated with docetaxel

Initially, the datasets of the four international, multicenter studies containing 1,479 atezolizumab-treated patients’ survival data (the baseline characteristics of these patients are summarized in Supplementary Table S1) were combined to identify 80 common BCT biomarkers that demonstrated correlations to PFS and OS in advanced NSCLC. Next, we removed the 62 BCT biomarkers that we identified from analyses of the survival data of 707 advanced NSCLC patients who underwent docetaxel treatment. Moreover, all biomarkers containing absolute cell counts were eliminated to avoid sampling-based systemic errors. Hence, 11 BCT biomarkers remained. Consecutively, we selected the cell ratios of NLR, PLR and LMR at 12 weeks on-treatment (NLR_T3, PLR_T3 and LMR_T3), and NMR at 6 and 12 weeks on-treatment (NMR_T2 and NMR_T3), for further analysis. Frequency distribution analysis and Wilcoxon signed-rank test of these four BCT biomarkers at pre-treatment (T1) showed no significant difference between the atezolizumab and docetaxel treatment groups in OAK and POPLAR studies (Supplementary Figure S1). Hence, we hypothesized that subsequent changes at T2 and T3 of these biomarkers might have more value (Supplementary Figure S2).

Next, we performed univariate Cox analysis by decile patient fractions at 10% intervals from 10% to 90% or by the quadrant percentiles of 25% and 75% for all five biomarkers in the combined datasets of atezolizumab-treated patients to calculate HRs for OS and PFS (Figure 2), respectively. NLR_T3 showed significant HR for all patient cutoffs examined in the atezolizumab-treated group and for both OS and PFS. PLR_T3 showed significant PFS HR for the >10% patient fractions and significant OS HR for all defined patient fractions. LMR_T3 showed significant but inconsistent PFS HR between the 25% and 80% patient fractions and significant OS HR for the >10% patient fraction. NMR_T2 showed significant PFS HR at the >20% patient fractions and significant OS HR at the >10% patient fractions. NMR_T3 showed significant PFS HR at the >10% patient fractions and significant OS HR for all patient fractions.

**Figure 2.**
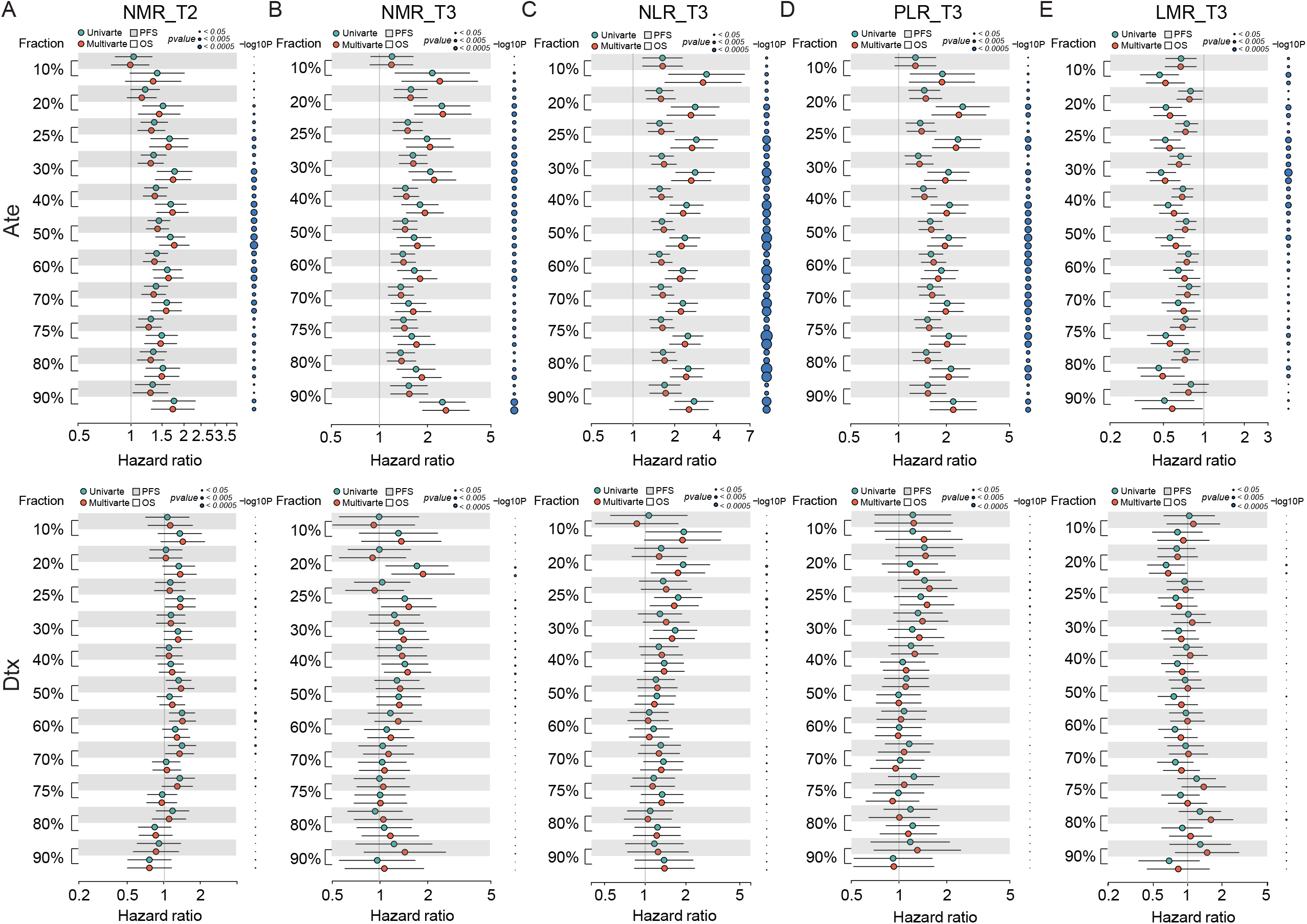
Forest plot of hazard ratio (HR) for overall survival (OS) and progression-free survival (PFS) of the BCT biomarkers (**A**) NMR_T2, (**B**) NMR_T3, (**C**) NLR_T3, (**D**) PLR_T3 and (**E**) LMR_T3 in decile patient fractions in the atezolizumab (Ate) or docetaxel (Dtx) treatment groups of the combined internal cohorts. Mean HRs for OS (white shade) or PFS (grey shade) under univariate (green) or multivariate (red) Cox analysis is indicated by the dots, the range of HR is indicated by the error bar of the forest plot; -log_10_ *p*-value of each calculated HR is indicated by the size of the blue dots adjacent to the forest plot.

On multivariate analysis, we initially screened for clinical factors that might confer to PFS and OS in atezolizumab-treated NSCLC patients (Supplementary Table S2). Similar to univariate analysis, we performed multivariate Cox analysis using the same patient fractions in the combined datasets for PFS and OS (Figure 2), respectively. NLR_T3, PLR_T3, NMR_T2 and NMR_T3 all depicted identical trends to univariate analysis. Alternatively, LMR_T3 showed significant PFS HR from 10% to 80% patient fractions and significant OS HR from 10% to 50% patient fractions. In contrast, all five BCT biomarkers showed non-significant HRs for both OS and PFS in the docetaxel-treated group (Figure 2). Collectively, these results suggested that LMR_T3 exhibited significant but inconsistent HRs as compared to the rest of the selected biomarkers. Furthermore, it is noteworthy that LMR_T3 displayed HR <1 whereas the other four biomarkers displayed HR > 1.

After that, we applied the univariate and multivariate Cox analysis with decile patient fractions to the cohort of atezolizumab-treated NSCLC patients in the four individual trials respectively (Supplementary Figure S3). In concordance to the joint analyses, all of the five biomarkers showed no significant HRs for both PFS and OS in the docetaxel treatment group. In contrast, positive results consistent to the combined cohort were obtained for all biomarkers in the BIRCH and OAK cohorts for both PFS and OS. This was also true of the POPLAR cohort, except for LMR_T3. However, in the FIR cohort none of the biomarkers demonstrated significant HRs for either PFS or OS, but this is most likely because of the small sample size ^22^. Consequently, absolute integer cutoff values were set for the combined cohort using the patient fractions of 25-50% for all five biomarkers to establish a BCTscore model. Application of these variables to univariate and multivariate Cox analysis of each trial’s cohort succeeded in narrowing the range of each biomarker’s integer cutoff values to uncover the significant range (Supplementary Figure S4). NLR_T3, PLR_T3, and NMR_T2 confirmed consistently significant PFS and OS HRs in the cohorts of BIRCH, OAK and POPLAR. In contrast, all cutoff values of LMR_T3 did not. Because LMR_T3 showed consistently poor prognostic power in all of the above outlined analyses, it was removed for the definition of BCTscore.

Hence, all combinations of the three selected BCT biomarkers, namely NLR_T3, PLR_T3 and NMR_T2, formed the 16 BCTscore candidates subjected to further optimization for clinical application (Supplementary Table S3).

### Optimization of BCT biomarker combinations to establish the BCTscore model

To establish the BCTscore model, the OAK study was used as our training cohort. Next, the BIRCH study was used as internal validation cohort 1, and the POPLAR combined with the FIR study as internal validation cohort 2. Univariate and multivariate Cox analysis demonstrated that all of the 16 BCTscore candidates demonstrated significant HRs in both OS and PFS, as well as RR for CB and ORR (Supplementary Figure S5). To further narrow down the BCTscore candidates, we performed ROC analysis for OS, PFS, CB and ORR. The BCTscore candidate 2 (BCTscore #2) was the only candidate that had good AUC for OS, PFS, CB and ORR in all of the three internal cohorts (Supplementary Table S4). Hence, BCTscore candidate 2, comprising of the BCT biomarkers of NLR and PLR at 12 weeks on-treatment (T3) and NMR at 6 weeks on-treatment (T2) with absolute cutoff values of NLR_T3 = 5, PLR_T3 = 180 and NMR_T2 = 6, respectively, was selected as the BCTscore model for NCSLC.

This BCTscore model displayed significant OS and PFS HRs in both univariate and multivariate Cox analysis in all of the three cohorts (Supplementary Figure S5A). The OAK cohort’s RR for CB (univariate = 0.60 (95% CI: 0.39-0.93), *P* = 0.024; multivariate = 0.56 (95% CI: 0.35-0.88), *P* = 0.014) (Supplementary Figure S5B) and ORR (univariate = 0.53 (95% CI: 0.31-0.91), *P* = 0.22; multivariate = 0.58 (95% CI: 0.37-0.88), *P* = 0.013) with BCTscore stratification (Supplementary Figure S5B) were good. The rate of CB (high-risk = 38%, low-risk = 51%) and ORR (high-risk = 17%, low-risk = 28%) of the low-risk atezolizumab-treated patients in the OAK cohort after BCTscore stratification (Supplementary Table S5) were also higher than the 48% CB and 14% ORR reported in the original study ^4^. Furthermore, survival analysis also showed that our newly identified BCTscore model presented significant difference in both OS and PFS between high-risk and low-risk patients in the atezolizumab-treated group (Figure 3). ROC analysis resulted in a BCTscore model that consistently exhibited better OS AUC in the OAK (AUC_12month_ = 0.696), BIRCH (AUC_12month_ = 0.672) and POPLAR+FIR studies (AUC_12month_ = 0.727) than that of each of the three single BCT biomarkers in these studies (Figure 4). However, the AUCs of the BCTscore model were lower than those of NLR_T3 for PFS (Supplementary Figure S6), CB (Supplementary Figure S7) and ORR (Supplementary Figure S7) in the OAK cohort, whereas the BCTscore model depicted better AUCs than the standalone BCT biomarkers for these survival indicators in the BIRCH and POPLAR+FIR cohorts.

**Figure 3.**
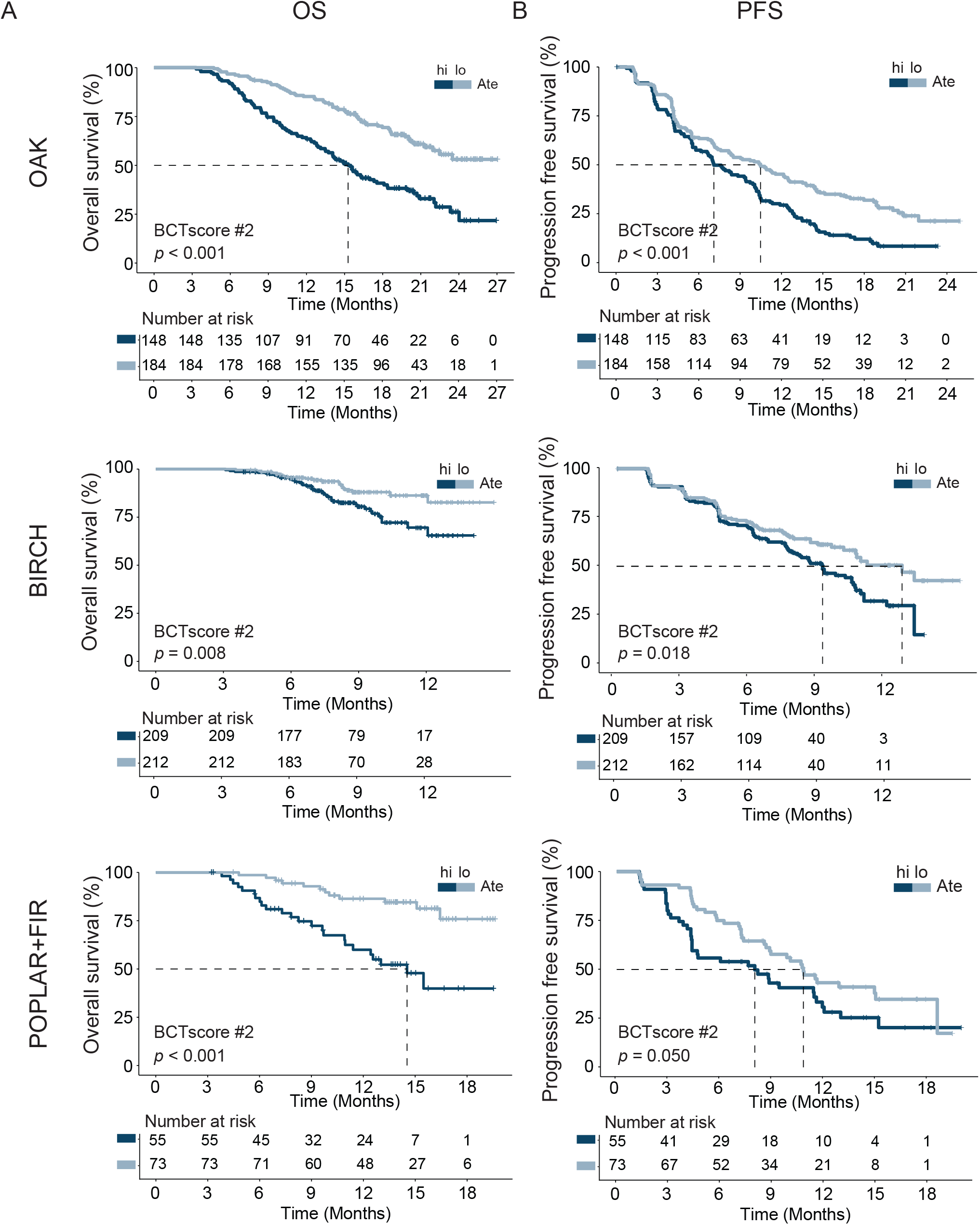
Kaplan-Meier analysis of (**A**) overall survival (OS) and (**B**) progression-free survival (PFS) between high-risk (hi) and low-risk (lo) patients, as defined with the identified BCTscore candidate 2 (BCTscore #2), treated with atezolizumab (Ate) of the training cohort (OAK) and the internal validation cohorts (BIRCH and POPLAR+FIR). The percentage of survival of high-risk (dark blue) and low-risk (light blue) patients is plotted against the time in months.

**Figure 4.**
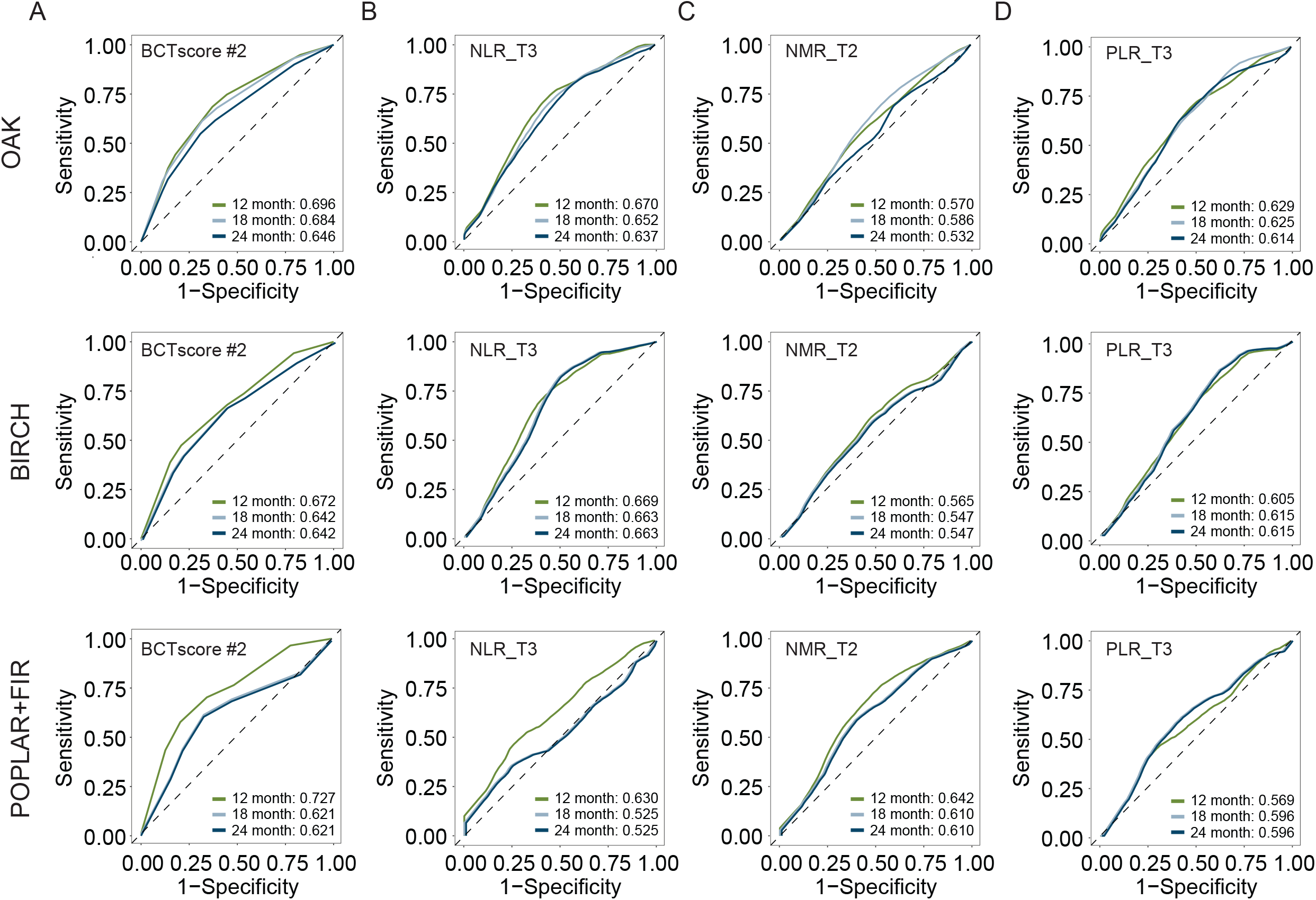
Time-dependent receiver-operating characteristic (ROC) analysis for overall survival (OS) to obtain the area under curve (AUC) of (**A**) BCTscore candidate 2 (BCTscore #2) and the BCT biomarkers (**B**) NLR_T3, (**C**) NMR_T2 and (**D**) PLR_T3 of the atezolizumab-treated patients of the training cohort (OAK) and the internal validation cohorts (BIRCH and POPLAR+FIR). Sensitivity is plotted against specificity.

### Investigation of the BCTscore model as a predictive biomarker

Lastly, in order to test if the developed BCTscore model could serve as a predictive biomarker, we performed survival analysis on the OAK and POPLAR RCTs. The Ate *vs* Dtx HRs of each BCT biomarker’s absolute cutoff value or decile fractionated BCTscore candidates above and below the cutoff were also calculated to determine if each BCTscore candidate was a predictive biomarker ^23^. Results showed that NLR_T3 presented significant PFS prognosis in the OAK study and may be prognostic of OS in the POPLAR study, whereas PLR_T3 and NMR_T2 achieved no significant results in both RCTs (Supplementary Table S6). On the other hand, all 16 BCTscore candidates had some predictive power in >75% fractions in the OAK and the POPLAR RCTs (Supplementary Table S7). In this regard, our newly developed BCTscore model is a strong predictive model specific to atezolizumab-treated NSCLC patients in comparison to docetaxel-treated patients for OS in the cohorts of OAK (BCTscore low-risk: HR _Ate vs Dtx_ = 1.54 (95% CI: 1.04-2.27), *P* = 0.036; high-risk: HR _Ate vs Dtx_ = 0.84 (95% CI: 0.62-1.12), *P* = 0.236) (Figure 5) and POPLAR (BCTscore low-risk: HR _Ate vs Dtx_ = 2.93 (95% CI: 1.21-7.10), *P* = 0.022; high-risk: HR _Ate vs Dtx_ = 0.56 (95% CI: 0.29-1.07), *P* = 0.078) (Supplementary Figure S8). In contrast, no significant difference was observed in PFS between the atezolizumab and docetaxel treatment groups in both the OAK (BCTscore low-risk: HR _Ate vs Dtx_ = 1.22 (95% CI: 0.85-1.75), *P* = 0.281; high-risk: HR _Ate vs Dtx_ = 0.79 (95% CI: 0.60-1.04), *P* = 0.090) (Supplementary Figure S9) and POPLAR studies (BCTscore low-risk: HR _Ate vs Dtx_ = 1.06 (95% CI: 0.50-2.24), *P* = 0.878; high-risk: HR _Ate vs Dtx_ = 0.87 (95% CI: 0.47-1.59), *P* = 0.645) (Supplementary Figure S10), in consistence to the findings of the two studies ^3, 4^. Similarly, analysis of the relative response rate suggested that our BCTscore model did not distinguish between the atezolizumab- and docetaxel-treated patients in both CB (OAK high-risk: Ate *vs* Dtx = 1.25, low-risk: Ate *vs* Dtx = 0.85; POPLAR high-risk: Ate *vs* Dtx = 0.67, low-risk: Ate *vs* Dtx = 0.95) and ORR (OAK high-risk: Ate *vs* Dtx = 0.96, low-risk: Ate *vs* Dtx = 0.95; POPLAR high-risk: Ate *vs* Dtx = 0.79, low-risk: Ate *vs* Dtx = 0.78) (Supplementary Table S8), reinforcing the fact that our newly defined BCTscore model is a predictive and prognostic biomarker particularly for OS.

**Figure 5.**
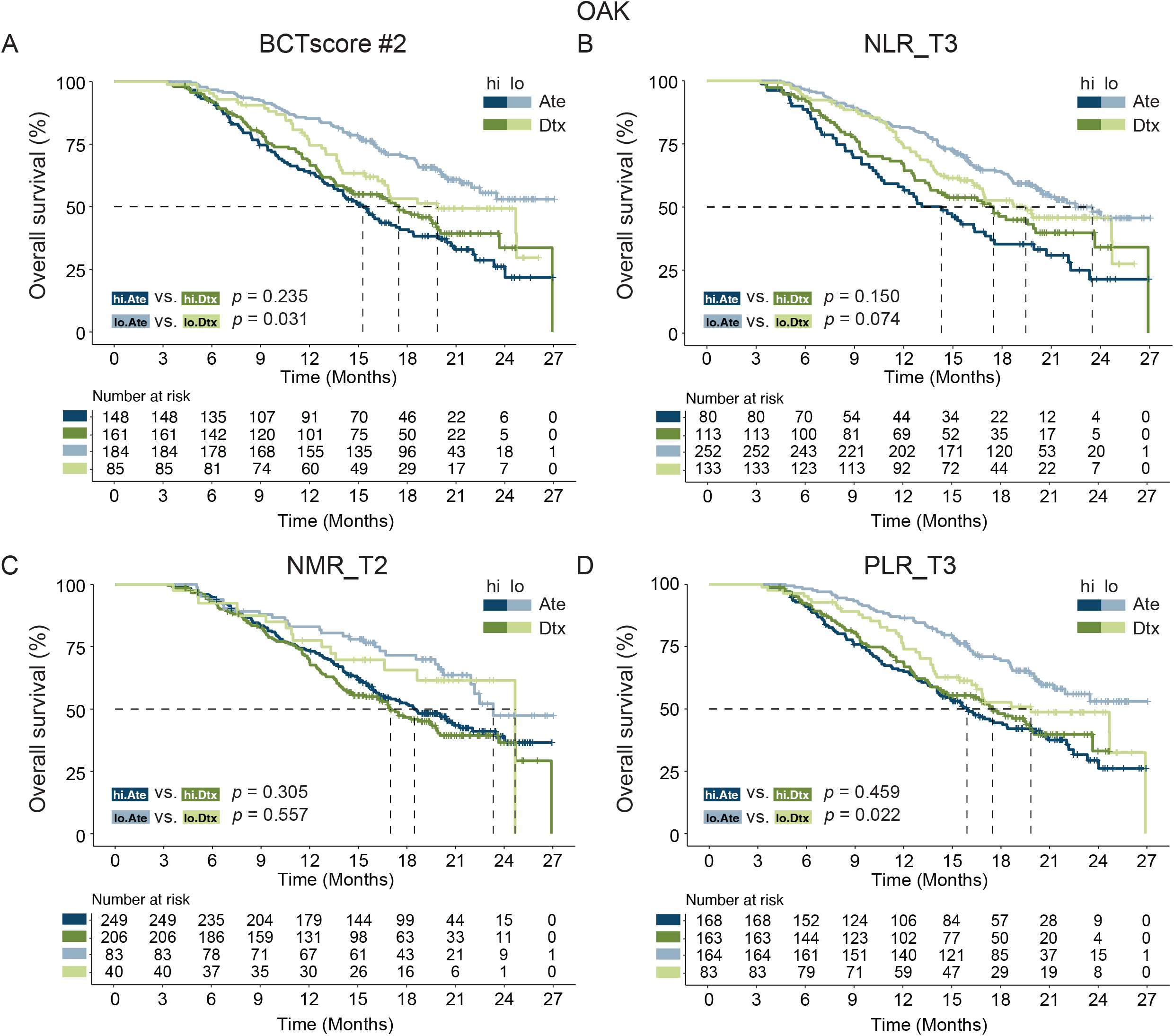
Kaplan-Meier analysis of overall survival (OS) for (**A**) BCTscore candidate 2 (BCTscore #2) and the BCT biomarkers (**B**) NLR_T3, (**C**) NMR_T2 and (**D**) PLR_T3 comparing atezolizumab (Ate)-treated patients (dark blue) against docetaxel (Dtx)-treated patients (dark green) in the high-risk (hi) group, and comparing between Ate-treated patients (light blue) against Dtx-treated patients (light green) in the low-risk (lo) group of the training cohort (OAK).

Taken together, these results clearly indicate that our BCTscore model can predict the efficacy of anti-PD-L1 ICI therapy of NSCLC patients in the clinical setting.

### BCTscore model validation in an external cohort

Because of the lack of real-world data on atezolizumab mono-treated NSCLC patients, we used an external validation cohort consisting of 168 advanced or relapsed NSCLC patients who received anti-PD-1-based combination therapy. Analysis of the external validation cohort by univariate Cox proportional hazard regression showed that our BCTscore model was relevant for OS (HR = 1.71 (95% CI: 1.00-2.91), *P* = 0.048), and a bit less for PFS (HR = 1.50 (95% CI: 0.97-2.32), *P* = 0.066) (Supplementary Figure S11A). Alternatively, the RR for CB (0.61 (95% CI: 0.30-1.20), *P* = 0.156) was comparable to that of the OAK cohort, whereas the low ORR (0.14 (95% CI: 0.02-0.55), *P* = 0.013) (Supplementary Figure S11B) was correlated to the lack of response in the original study ^21^. The Kaplan-Meier curves showed that our BCTscore model had a slight, but non-significant, contribution for both OS (HR = 0.72 (95% CI: 0.44-1.16), *P* = 0.169) (Supplementary Figure S11C) and PFS (HR = 0.76 (95% CI: 0.52-1.11), *P* = 0.157) (Supplementary Figure S11D). ROC analysis showed tolerable AUC for OS (AUC_12month_ = 0.557) (Supplementary Figure S11E), PFS (AUC_12month_ = 0.526) (Supplementary Figure S11F), CB (AUC_12month_ = 0.434) (Supplementary Figure S11G) and ORR (AUC_12month_ = 0.746) (Supplementary Figure S11H).

## Discussion

In spite of the success of ICI therapy in NSCLC treatment, robust prediction of treatment response remains one of the biggest challenges ^24^. BCT routinely performed test in the clinic that provides an unbiased overview of the immune landscape for patient stratification and longitudinal ICI efficacy assessment without the need for specialized analysis. This study showed that the BCTscore model serves as both a strong prognostic and predictive biomarkers of ICI efficacy. The strengths of this study are manifold. First, the OS AUCs of our newly identified BCTscore model surpassed that of PD-L1 ^25^ and TMB^26, 27^, both of which can only be applied by more invasive tissue biopsy procedures. Next, an important feature of our analyses is easily obtainable longitudinal data. We presume that the immune landscape alters during treatment, so each time point signifies a discrete event ^28^. Simultaneously, we removed any BCT biomarker at any timepoint that is correlated to patient survival after docetaxel treatment during initial biomarker screening, thus selecting atezolizumab-specific BCT biomarkers. Our assumption is that BCT biomarkers typical to cancer prognosis regardless of biological mechanism will show significant HR in the docetaxel patient group and hence should be removed from subsequent analyses. Hence, prognostic biomarkers were ruled out using our approach.

Furthermore, delays in immune response have been frequently observed during ICI therapy ^29, 30^. That is why later time points are hypothesized to have better indicative power as compared to earlier time points. Nevertheless, the biological nature of the BCT biomarker still holds the key to a successful predictive biomarker. For example, both NMR_T2 and NMR_T3 depicted significant HRs in our preliminary analysis. However, NMR_T2 showed better results in the survival analysis of the OAK and POPLAR studies than NMR_T3, whereas NLR_T3 and PLR_T3 displayed good results in the same analysis. This result demonstrated that after the number of neutrophils increased at 6 weeks on-treatment (T2), subsequent increase had little impact on disease prognosis and prediction; instead, the reduction of lymphocytes and increase in platelet numbers at 12 weeks on-treatment (T3) come into play. No prior studies have examined this, to our understanding, in the context of ICI, and the biological mechanism on the temporal changes of the immune landscape during ICI treatment remains elusive. Additionally, we picked IC ratios over absolute cell numbers to avoid systemic errors during blood sampling and sample analysis performed in different medical centers. The cell ratios used for our analysis were selected based on cell lineage and published data. For example, NMR and NLR were picked to distinguish changes in the neutrophil population against the IC lineages of monocytes and lymphocytes. It is known that lymphocytes are directly involved in tumor killing, whereas monocytes have more diverse biological roles. Indeed, the fact that NLR outperforms all other cell ratios as a single BCT biomarker reinforces previous observations that increasing neutrophil numbers and decreasing lymphocyte numbers result in poor cancer prognosis ^31, 32^. Alternatively, the poor correlation between LMR and survival supports the hypothesis that neutrophils, but not the entire monocyte population, contribute to cancer prognosis. Next, the improvement in prognostic ability by combining multiple IC ratios suggests that many factors play a role in ICI treatment; for instance, neutrophils were shown to promote tumor metastases ^33, 34^; platelets present antigens to trigger immune evasion ^35^. We deduce that future immunological studies will deepen our knowledge of the correlation between the immune landscape and ICI treatment success to unravel more effective and accurate biomarkers ^36^.

Finally, analysis of the four international, multi-center clinical trials consisting of 1,480 NSCLC patients treated with atezolizumab and validation in 151 patients treated with anti-PD-1 combination therapy provides strong statistical evidence to support our findings. We observed statistically significant associations for OS, PFS, ORR, and CB with powerful diagnostic abilities, suggesting that the newly defined BCTscore has prognostic and predictive value in the context of anti-PD-L1/PD-1 ICI therapy. Nevertheless, because the mechanistic role of the PD-1/PD-L1 immune biomarkers is conserved in tumor recognition by T cells, application of our model to other ICI therapies, such as anti-CTLA-4 therapy, remains to be tested.

A limitation of our study is the lack of TMB measurements in our dataset that would enable direct comparisons between the predictive power of our newly defined BCTscore model and TMB. We were restricted in the validation of our results because the external validation cohort was obtained with combination therapy with anti-PD-1 drugs due to the shortage of patients treated by atezolizumab alone. Although our BCTscore model did not reach statistical significance to predict OS and PFS in the used external validation cohort, the significant HR and tolerable OS AUC strongly suggest that this BCT biomarker combination is clinically relevant. Hence, optimization of the absolute cutoff values of each BCT biomarker will be investigated for different ICI therapies as well as combination therapies in future studies.

In summary, we demonstrated for the first time via a post-hoc analyses of four clinical trials the predictive value of longitudinal blood cell count ratio for NSCLC patients treated with ICI. Together, the training, internal, and external validation cohorts proved that the BCTscore combination of NLR at 12 weeks, PLR at 12 weeks, and NMR at 6 weeks provides prognostic and predictive information without the need to re-biopsy patients undergoing anti-PD-L1/PD-1 monotherapy. Future studies utilizing our BCTscore model may demonstrate its broader versatility as a prognostic and predictive biomarker in all lung cancer patients undergoing ICI treatment.

## Data Availability

We would like to thank all of the patients, investigators and staff involved in the FIR, BIRCH, POPLAR and OAK studies who released and shared their data. This publication is based on research using data from data contributors, Roche, that has been made available through Vivli, Inc. (Data Request ID: 5935; Lead Investigator: Dr. Jian-Guo Zhou). Vivli has not contributed to or approved, and is not in any way responsible for, the contents of this publication.

## Author Contributions

Conceptualization: JGZ, AHW, JGL, HM, USG, SHJ, DSS; methodology: JGZ, AHW, JGL, FYT; validation: YZC, BS, RF, MH, HM, USG, JGZ; data curation: JGZ, AHW, GS, SSH, HM, SRC, USG; writing—original draft preparation: AHW, HTW, JGZ, HTW; writing—review and editing: USG, HM, BF, SRC, DSS, MH; visualization: JGZ, HTW; supervision: HM, USG, JGZ, DSS; project administration: JGZ, HM, USG, RHW; funding acquisition: JGZ, SHJ, AHW. All authors have read and agreed to the published version of the manuscript.

## Acknowledgments

We would like to thank all of the patients, investigators and staff involved in the FIR, BIRCH, POPLAR and OAK studies who released and shared their data. This publication is based on research using data from data contributors, Roche, that has been made available through Vivli, Inc. (Data Request ID: 5935; Lead Investigator: Dr. Jian-Guo Zhou). Vivli has not contributed to or approved, and is not in any way responsible for, the contents of this publication. The present work was performed by Jian-Guo Zhou in partial fulfilment of the requirements for containing the degree “Dr. rer. biol. hum”.

## Patient consent for publication

Not required.

## Ethics approval

The studies were approved by the respective national ethics committees and institutional review boards and written informed consent was obtained from all patients.

## Provenance and peer review

Not commissioned; externally peer reviewed.

## Data availability statement

Data are available upon reasonable request from lead Investigator: Jian-Guo Zhou.

## SUPPLEMENTARY FIGURE LEGENDS

**Supplementary Figure S1**. Density plots of the ratio distribution of the BCT biomarkers (**A**) NLR, (**B**) PLR, (**C**) NMR and (**D**) LMR at baseline (T1) in the atezolizumab (Ate) and docetaxel (Dtx) treatment groups of the internal randomized controlled trials (OAK+POPLAR), accompanied by and (**E**) the comparison chart and *p*-values calculated by the Wilcoxon signed-rank test. Ratio density is plotted against the absolute ratio (except log_10_ ratio for PLR).

**Supplementary Figure S2**. Density plots of the ratio distribution of the BCT biomarkers (**A**) NLR, (**B**) PLR, (**C**) NMR and (**D**) LMR at baseline (T1), 6 weeks (T2) and 12 weeks (T3) on-treatment in the atezolizumab (Ate) and docetaxel (Dtx) treatment groups of the combined internal cohorts. Ratio density is plotted against the absolute ratio (except log_10_ ratio for PLR).

**Supplementary Figure S3**. Forest plot of hazard ratio (HR) for overall survival (OS) and progression-free survival (PFS) of the BCT biomarkers (**A**) NMR_T2, (**B**) NMR_T3, (**C**) NLR_T3, (**D**) PLR_T3 and (**E**) LMR_T3 in decile patient fractions in the atezolizumab-treated patients of each internal cohort (OAK, BIRCH, POPLAR, FIR). Mean HRs for OS (white shade) or PFS (grey shade) under univariate (green) or multivariate (red) Cox analysis is indicated by the dots, the range of HR is indicated by the error bar of the forest plot; -log_10_ *p*-value of each calculated HR is indicated by the size of the blue dots adjacent to the forest plot.

**Supplementary Figure S4**. Forest plot of hazard ratio (HR) for overall survival (OS) and progression-free survival (PFS) of the BCT biomarkers (**A**) NMR_T2, (**B**) NMR_T3, (**C**) NLR_T3, (**D**) PLR_T3 and (**E**) LMR_T3 at different absolute cutoff values in the atezolizumab (Ate) or docetaxel (Dtx) treatment groups of the combined internal cohorts. Mean HRs for OS (white shade) or PFS (grey shade) under univariate (green) or multivariate (red) Cox analysis is indicated by the dots, the range of HR is indicated by the error bar of the forest plot; -log_10_ *p*-value of each calculated HR is indicated by the size of the blue dots adjacent to the forest plot.

**Supplementary Figure S5**. Forest plot of (**A**) hazard ratio (HR) for overall survival (OS) and progression-free survival (PFS) and (**B**) relative risk (RR) for clinical benefit (CB) and objective response rate (ORR) of the BCTscore candidates in the atezolizumab-treated patients of the training cohort (OAK) and the internal validation cohorts (BIRCH and POPLAR+FIR). Mean HRs for OS (white shade) or PFS (grey shade) or RRs for CB (white shade) or ORR (grey shade) under univariate (green) or multivariate (red) Cox analysis is indicated by the dots, the range of HR or RR is indicated by the error bar of the forest plot; -log_10_ *p*-value of each calculated HR or RR is indicated by the size of the blue dots adjacent to the forest plot.

**Supplementary Figure S6**. Time-dependent receiver-operating characteristic (ROC) analysis for progression-free survival (PFS) to obtain the area under curve (AUC) of (**A**) BCTscore candidate 2 (BCTscore #2) and the BCT biomarkers (**B**) NLR_T3, (**C**) NMR_T2 and (**D**) PLR_T3 of the atezolizumab-treated patients of the training cohort (OAK) and the internal validation cohorts (BIRCH and POPLAR+FIR). Sensitivity is plotted against specificity.

**Supplementary Figure S7**. Receiver-operating characteristic (ROC) analysis for (**A**) clinical benefit (CB) and (**B**) objective response rate (ORR) to obtain the area under curve (AUC) of BCTscore candidate 2 (BCTscore #2), compared to the AUCs of the BCT biomarkers NLR_T3, NMR_T2 and PLR_T3 of the atezolizumab-treated patients of the training cohort (OAK) and the internal validation cohorts (BIRCH and POPLAR+FIR). Sensitivity is plotted against specificity.

**Supplementary Figure S8**. Kaplan-Meier analysis of overall survival (OS) for (**A**) BCTscore candidate 2 (BCTscore #2) and the BCT biomarkers (**B**) NLR_T3, (**C**) NMR_T2 and (**D**) PLR_T3 comparing between atezolizumab (Ate)-treated patients (dark blue) against docetaxel (Dtx)-treated patients (dark green) in the high-risk (hi) group, and comparing between Ate-treated patients (light blue) against Dtx-treated patients (light green) in the low-risk (lo) group of the internal validation cohort (POPLAR).

**Supplementary Figure S9**. Kaplan-Meier analysis of progression-free survival (PFS) for (**A**) BCTscore candidate 2 (BCTscore #2) and the BCT biomarkers (**B**) NLR_T3, (**C**) NMR_T2 and (**D**) PLR_T3 comparing between atezolizumab (Ate)-treated patients (dark blue) against docetaxel (Dtx)-treated patients (dark green) in the high-risk (hi) group, and comparing between Ate-treated patients (light blue) against Dtx-treated patients (light green) in the low-risk (lo) group of the training cohort (OAK).

**Supplementary Figure S10**. Kaplan-Meier analysis of progression-free survival (PFS) for (**A**) BCTscore candidate 2 (BCTscore #2) and the BCT biomarkers (**B**) NLR_T3, (**C**) NMR_T2 and (**D**) PLR_T3 comparing between atezolizumab (Ate)-treated patients (dark blue) against docetaxel (Dtx)-treated patients (dark green) in the high-risk (hi) group, and comparing between Ate-treated patients (light blue) against Dtx-treated patients (light green) in the low-risk (lo) group of the internal validation cohort (POPLAR).

**Supplementary Figure S11**. Forest plot of (**A**) hazard ratio (HR) for overall survival (OS) and progression-free survival (PFS) and (**B**) relative risk (RR) for clinical benefit (CB) and objective response rate (ORR) of the BCTscore candidates in the patients of the external validation cohort (JCH_ICI). Mean HRs for OS (white shade) or PFS (grey shade) or RRs for CB (white shade) or ORR (grey shade) under univariate (green) or multivariate (red) Cox analysis is indicated by the dots, the range of HR or RR is indicated by the error bar of the forest plot; -log_10_ *p*-value of each calculated HR or RR is indicated by the size of the blue dots adjacent to the forest plot. Kaplan-Meier analysis of (**C**) OS and (**D**) PFS between high-risk (hi) and low-risk (lo) patients of the external validation cohort (JCH_ICI). The percentage of survival of high-risk (dark blue) and low-risk (light blue) patients is plotted against the time in months. Time-dependent receiver-operating characteristic (ROC) analysis for overall survival (OS) to obtain the area under curve (AUC) for (**E**) OS, (**F**) PFS, (**G**) CB and (**H**) ORR of BCTscore candidate 2 (BCTscore #2) of the external validation cohort (JCH_ICI). Sensitivity is plotted against specificity.

## Footnotes

† PD-1, programmed cell death protein 1

‡ CD38 and CD39, clusters of differentiation 38 and 39

