## Supplementary figures and images for "Definition of a new blood cell count (BCT) score for early survival prediction for non-small cell lung cancer patients treated with atezolizumab: Integrated analysis of 4 multicenter clinical trials"

### Suppl Fig1

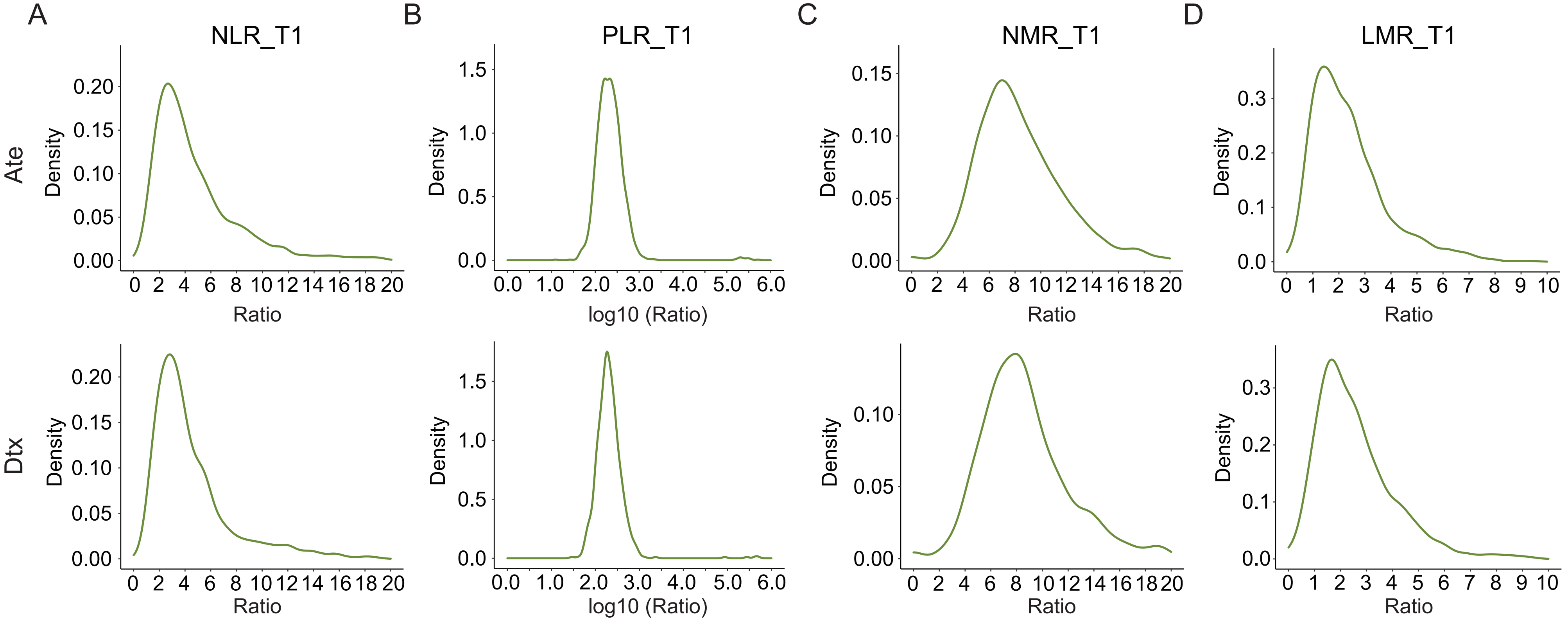


E

| | Ate | Dtx | | |
|---|---|---|---|---|
| BCT Biomarker | Mean Ratio | | Width | *p*-value |
| NLR_T1 | 5 | 5 | 270603 | 0.363 |
| PLR_T1 | 1441 | 2054 | 269252 | 0.488 |
| NMR_T1 | 24 | 10 | 250292 | 0.334 |
| LMR_T1 | 5 | 3 | 246724 | 0.143 |

### Suppl Fig2

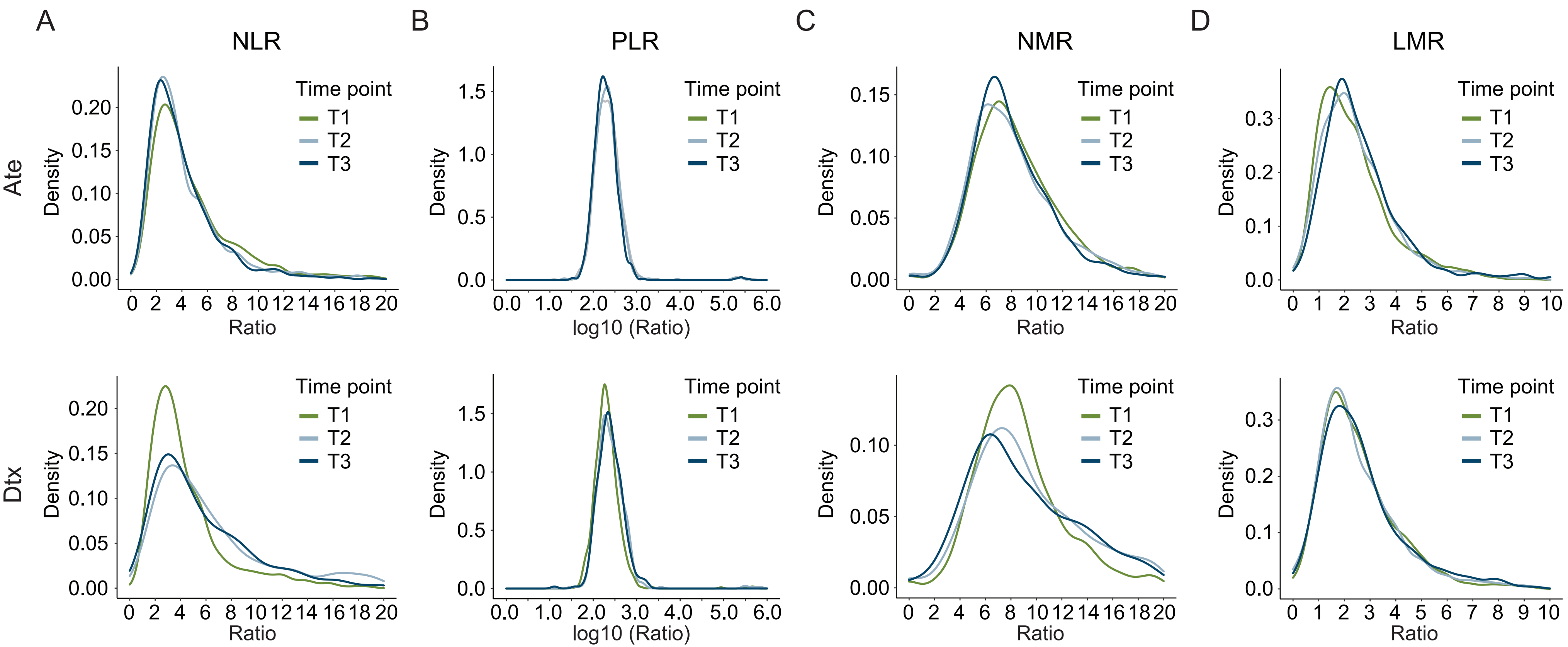

### Suppl Fig3

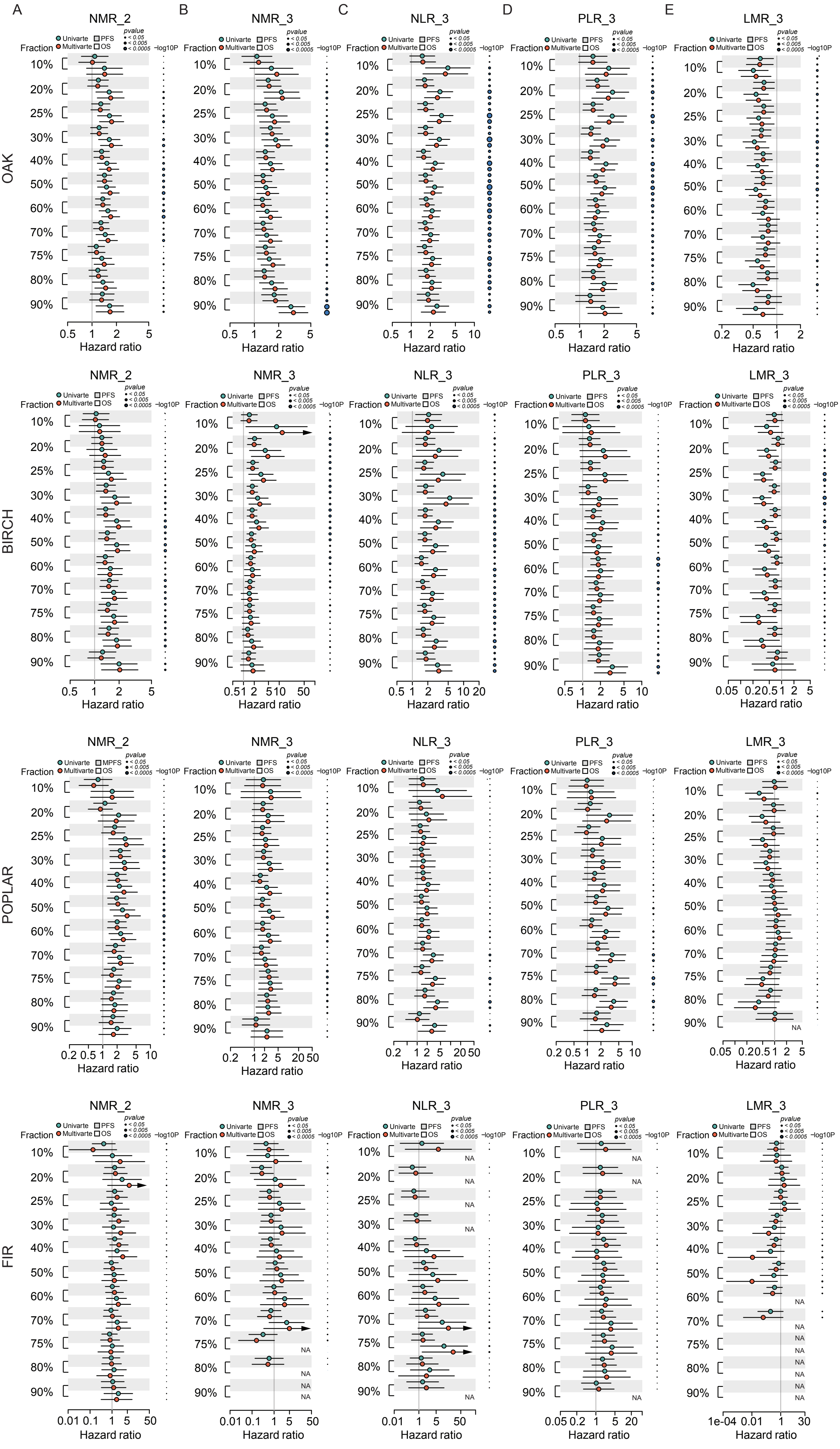

### Suppl Fig4

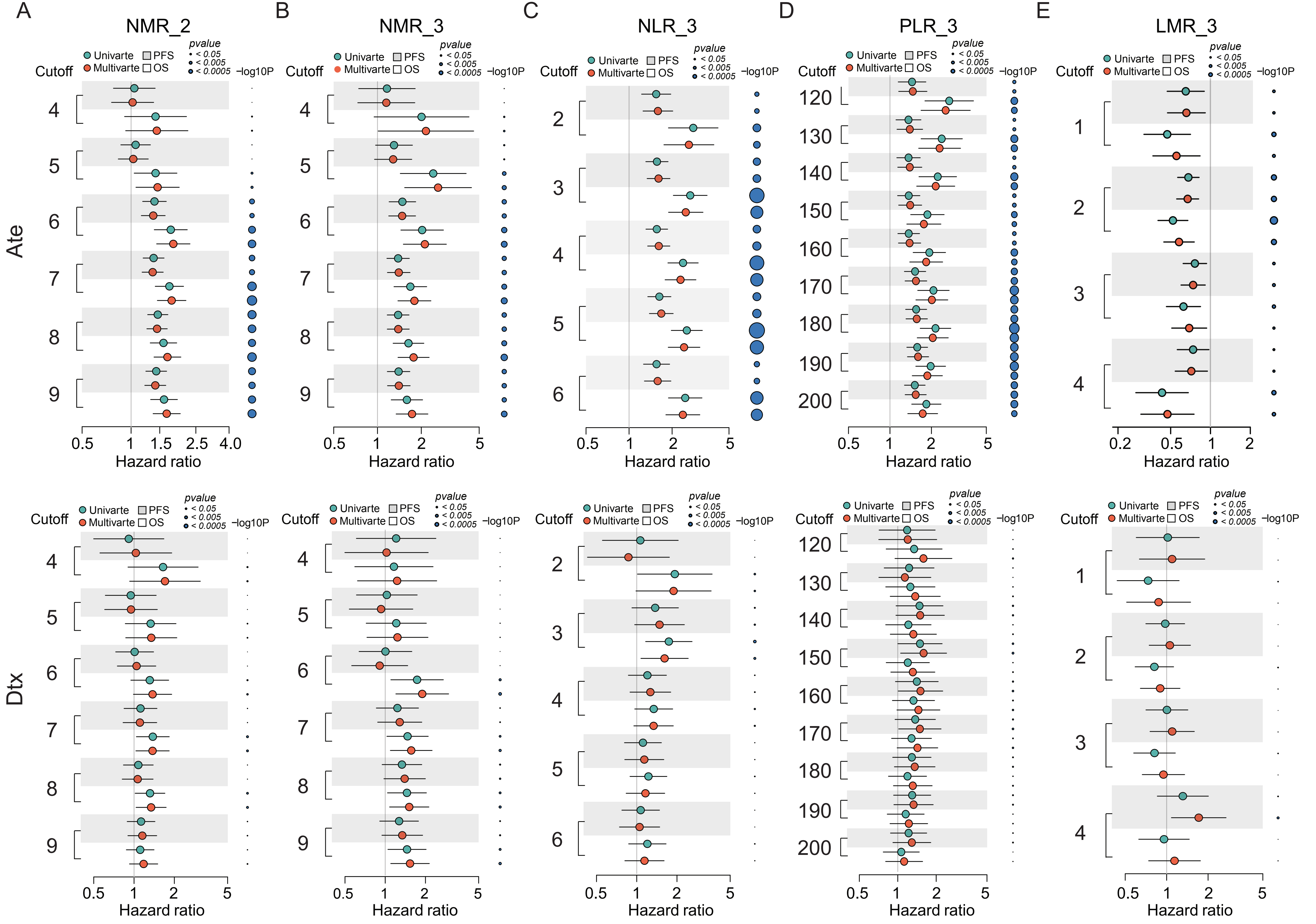

### Suppl Fig5

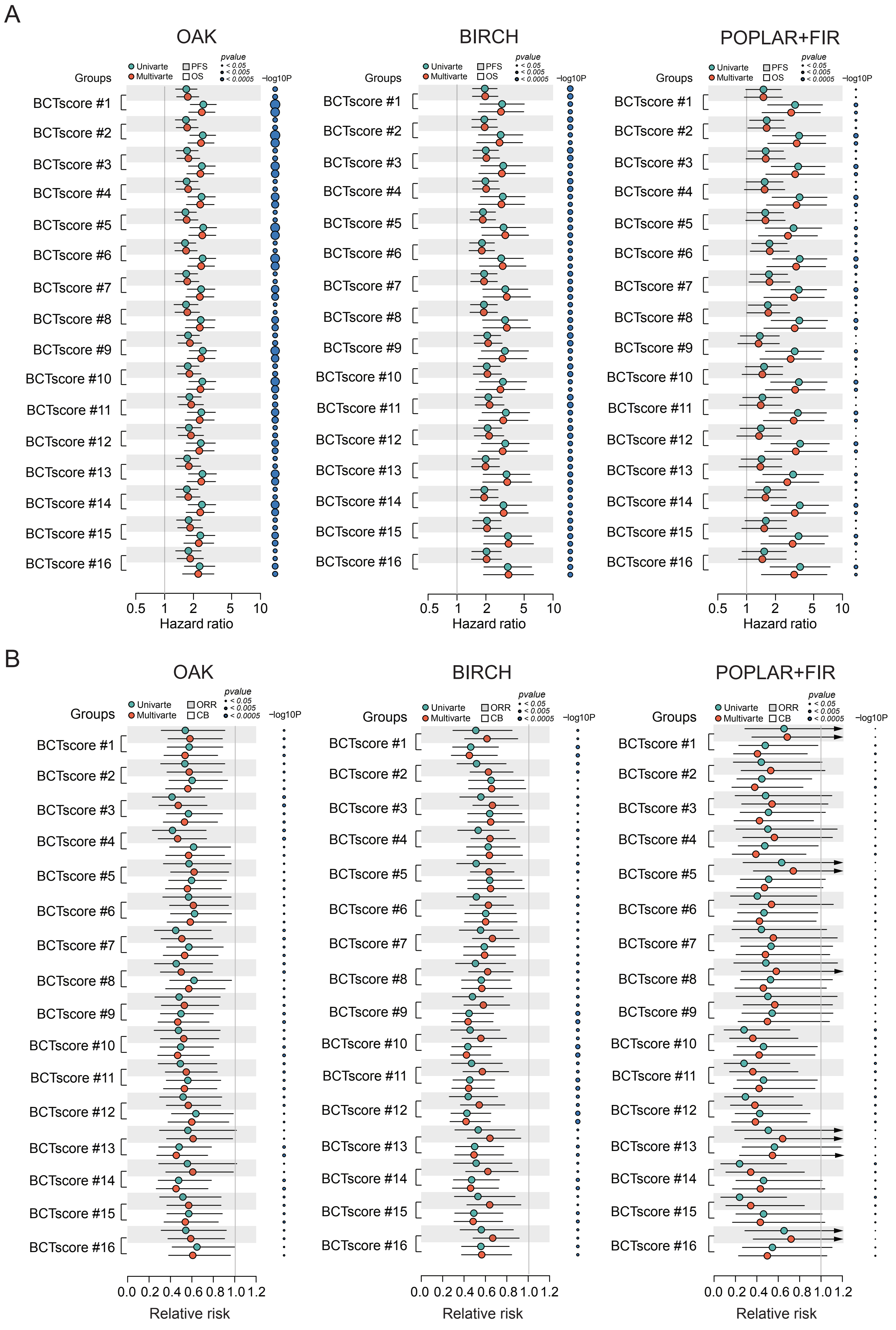

### Suppl Fig6

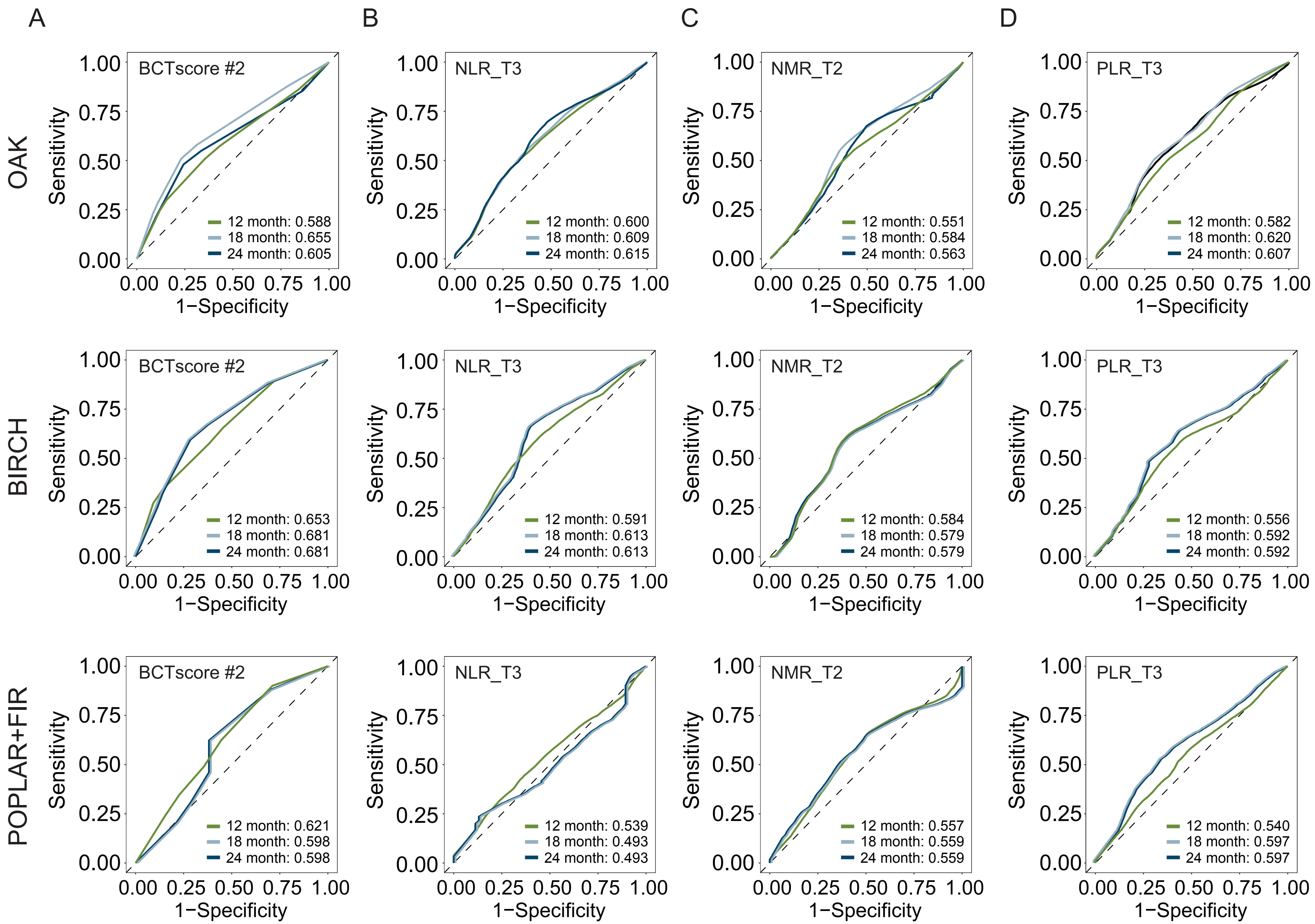

### Suppl Fig7

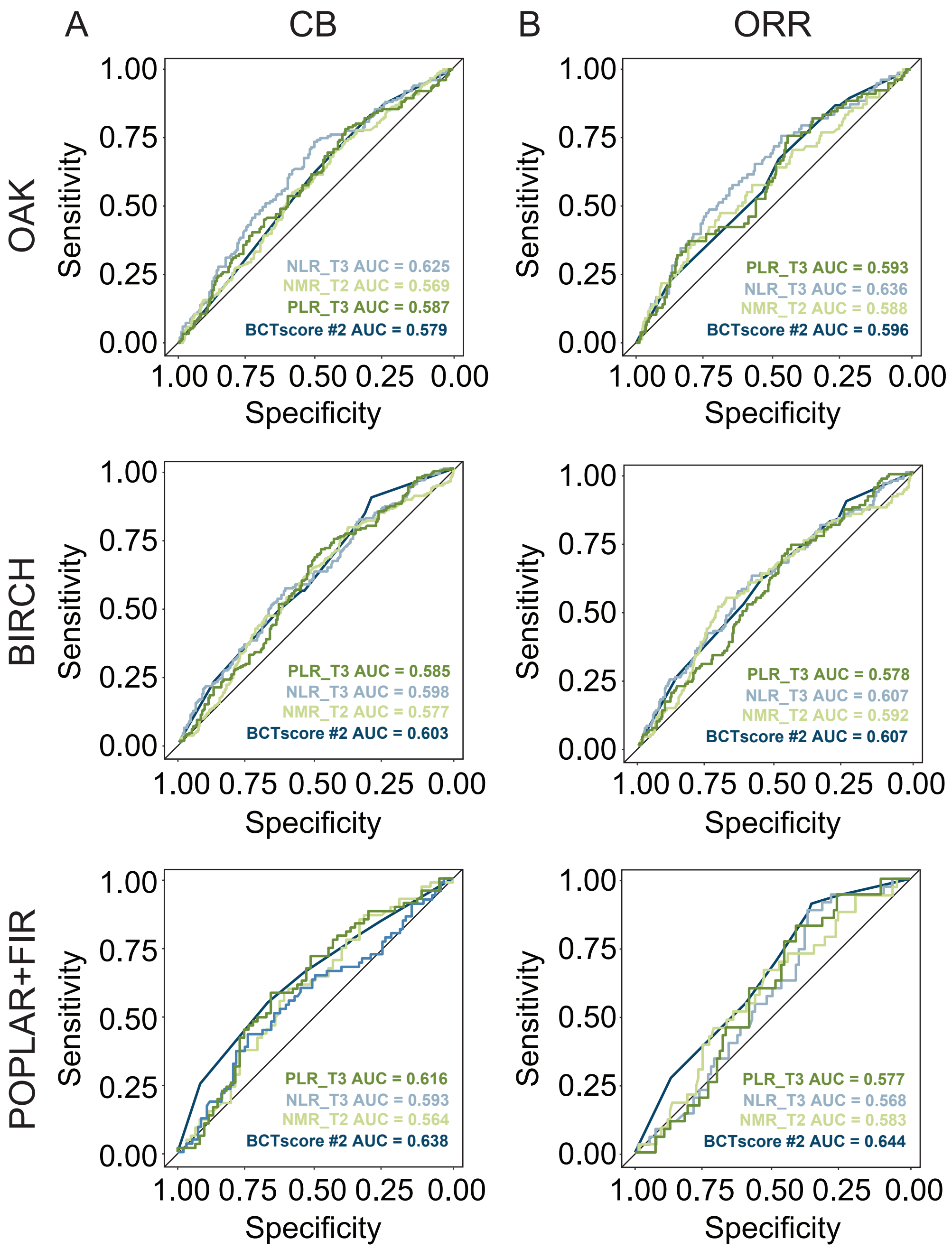

### Suppl Fig8

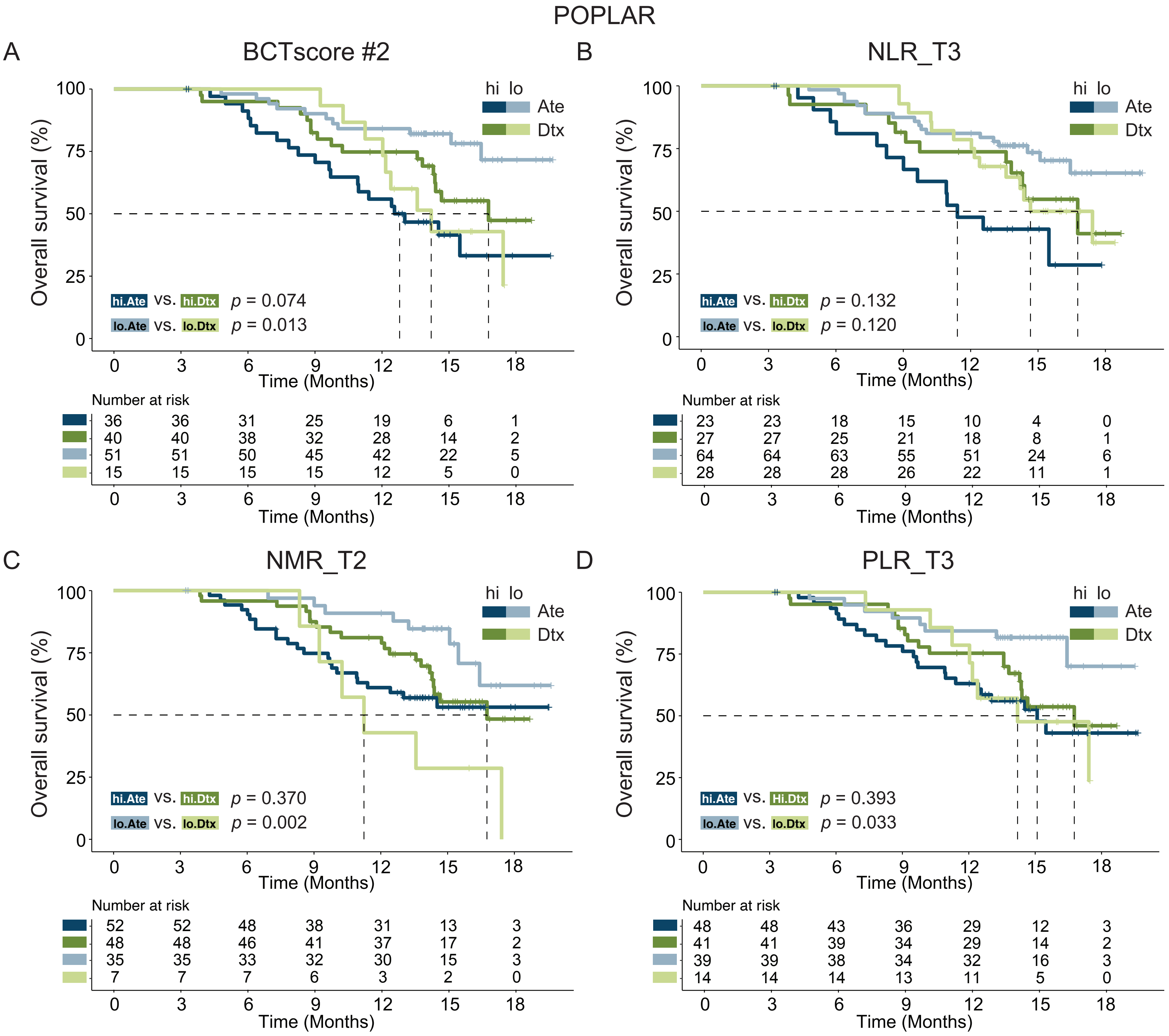

### Suppl Fig9

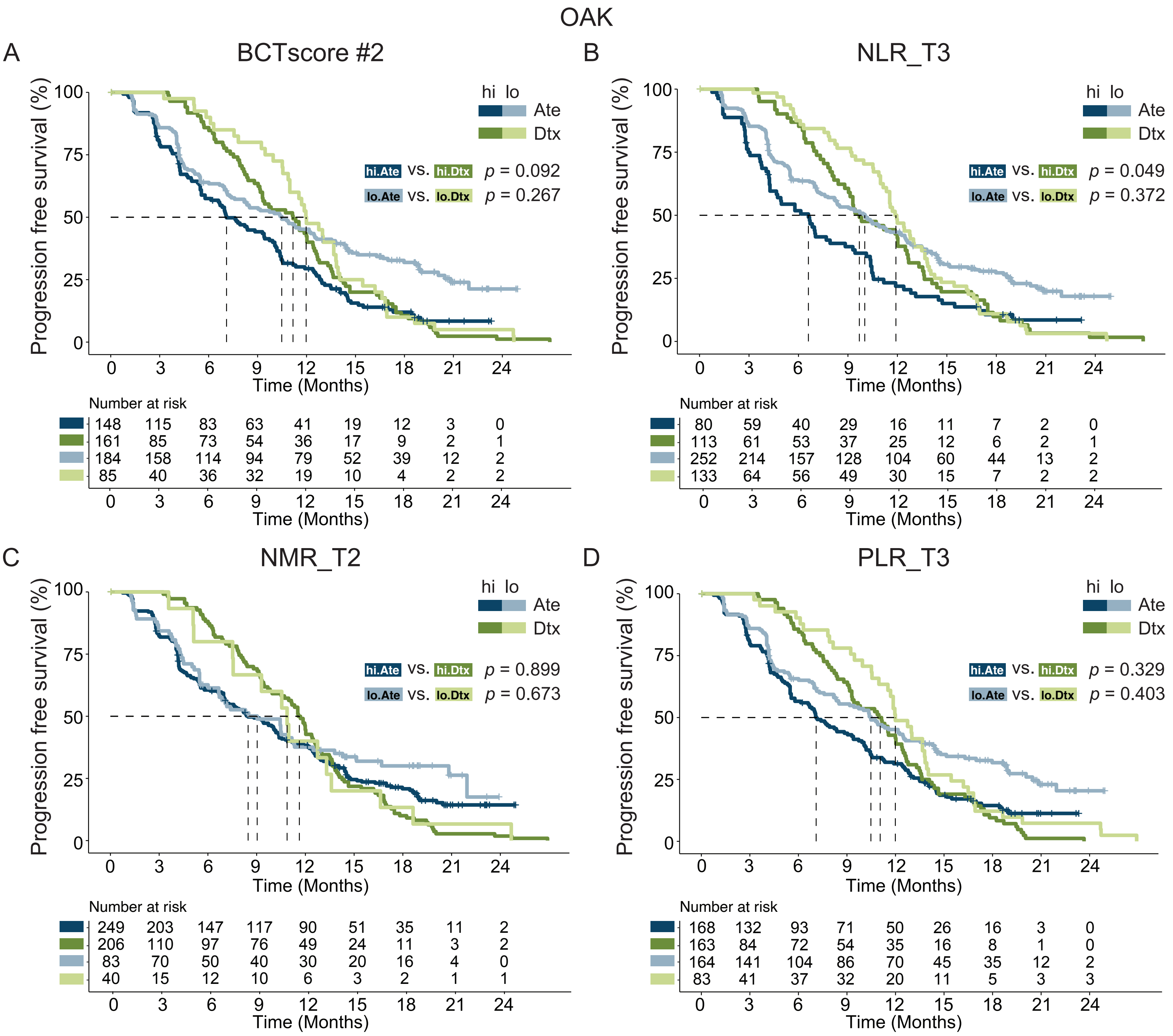

### Suppl Fig10

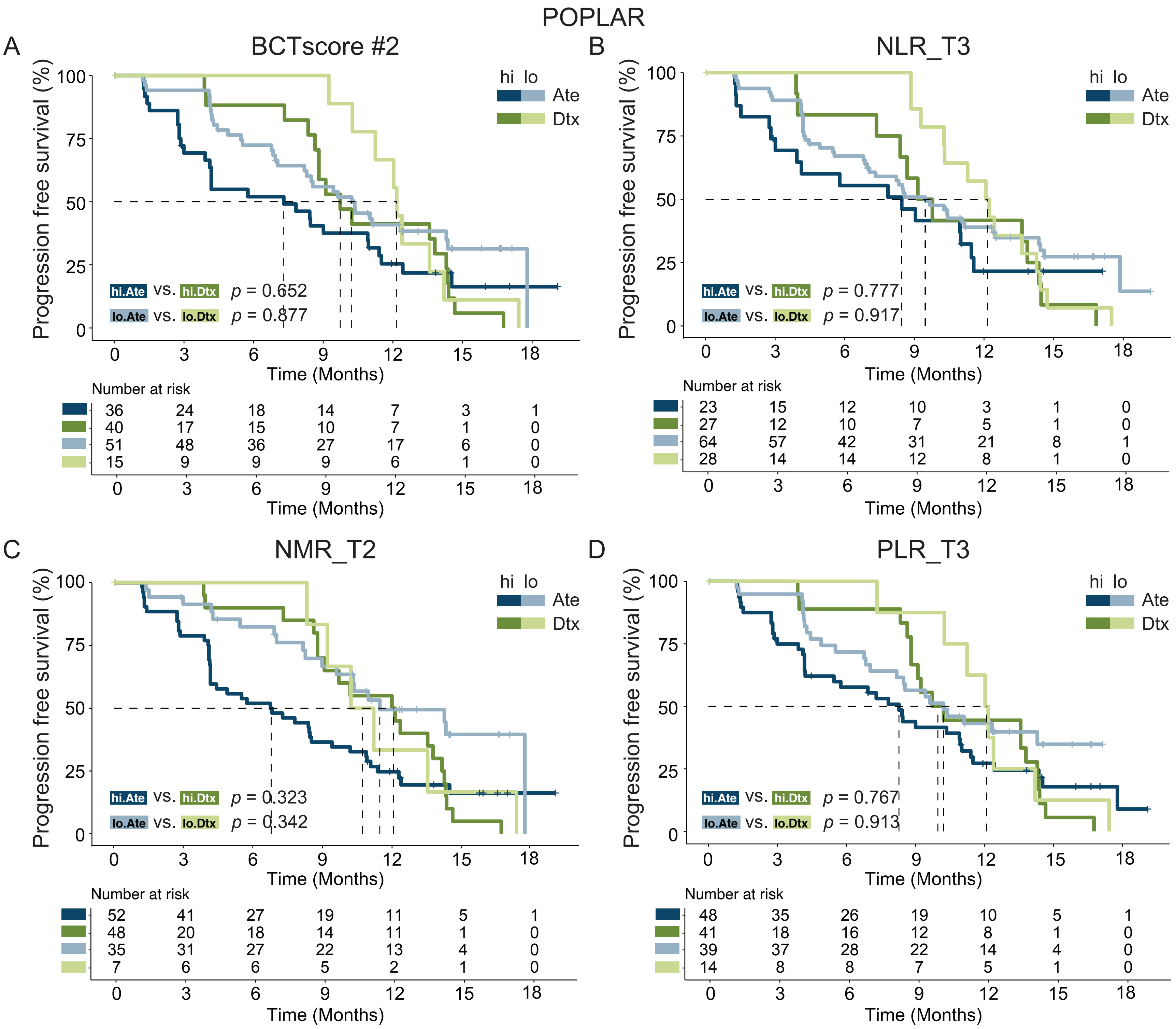

### Suppl Fig11

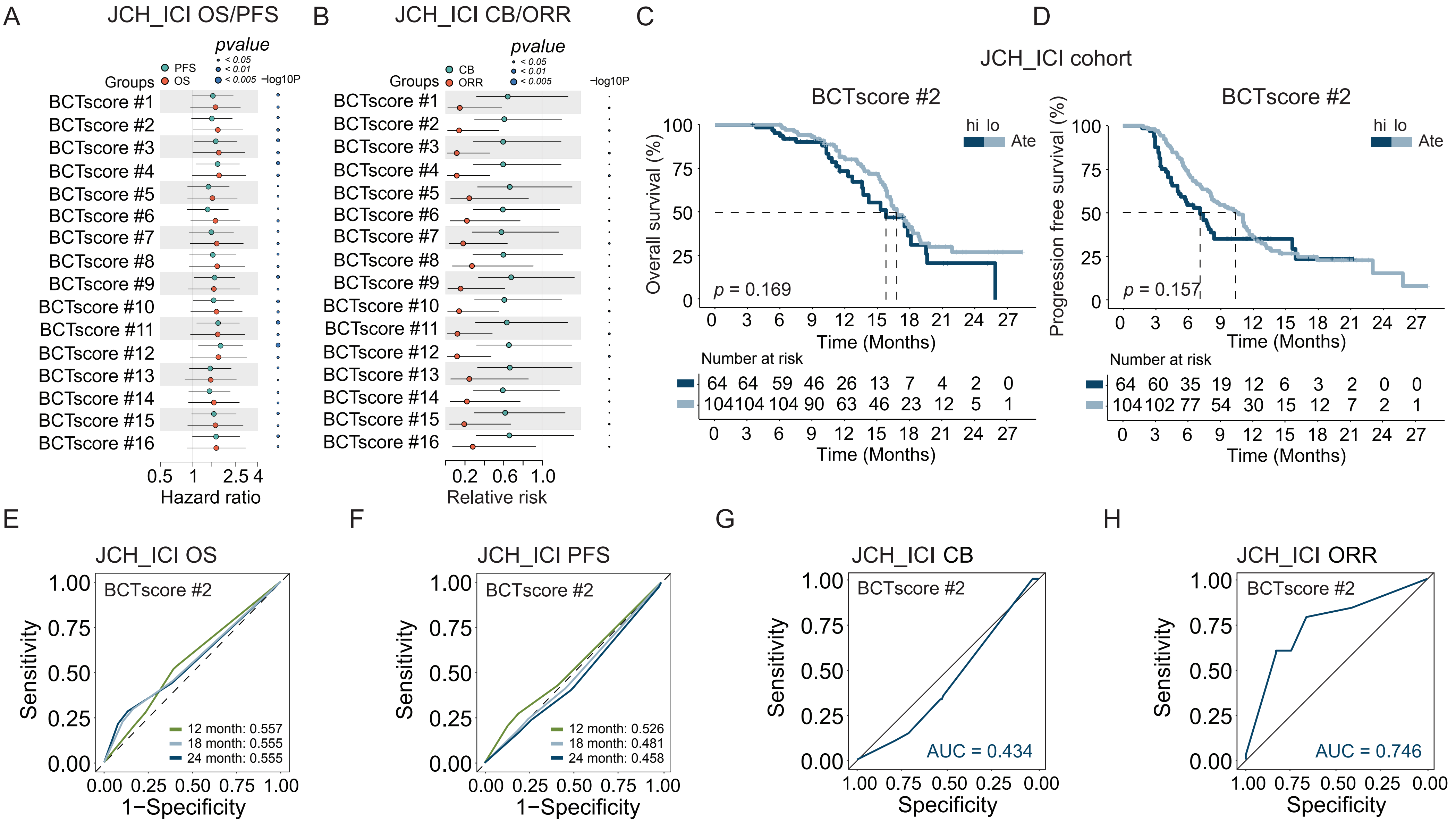
